# Application of concise machine learning to construct accurate and interpretable EHR computable phenotypes

**DOI:** 10.1101/2020.12.12.20248005

**Authors:** William La Cava, Paul C Lee, Imran Ajmal, Xiruo Ding, Priyanka Solanki, Jordana B Cohen, Jason H Moore, Daniel S Herman

## Abstract

**Objective:** Electronic health records (EHRs) can improve patient care by enabling systematic identification of patients for targeted decision support. But, this requires scalable learning of computable phenotypes. To this end, we developed the feature engineering automation tool (FEAT) and assessed it in targeting screening for the underdiagnosed, under-treated disease primary aldosteronism.

**Materials and Methods:** We selected 1,199 subjects receiving longitudinal care in a large health system and classified them for hypertension (N=608), hypertension with unexplained hypokalemia (N=172), and apparent treatment-resistant hypertension (N=176) by chart review. We derived 331 features from EHR encounters, diagnoses, laboratories, medications, vitals, and notes. We modified FEAT to encourage model parsimony and compared its models’ performance and interpretability to those of expert-curated heuristics and conventional machine learning.

**Results:** FEAT models trained to replicate expert-curated heuristics had higher area under the precision-recall curve (AUPRC) than all other models (*p* < 0.001) except random forests and were smaller than all other models (*p* < 1e-6) except decision trees. FEAT models trained to predict chart review phenotypes exhibited similar AUPRC to penalized logistic regression while being simpler than all other models (p < 1e-6). For treatment-resistant hypertension, FEAT learned a six-feature, clinically intuitive model that demonstrated a positive predictive value of 0.70 and sensitivity of 0.62 in held-out testing data.

**Discussion:** FEAT learns computable phenotypes that approach the performance of expert-curated heuristics and conventional machine learning without sacrificing interpretability.

**Conclusion:** By constructing accurate and interpretable computable phenotypes at scale, FEAT has the potential to facilitate systematic clinical decision support.

## INTRODUCTION

The adoption of electronic health records (EHRs) is transforming medicine by aiding clinical decision making and facilitating translational research.[1,2] In order to leverage EHR data, practitioners must first define rules or algorithms known as *computable phenotypes* that identify patient cohorts with certain characteristics of interest.[3–5] While there have been significant advances in creating and standardizing computable phenotypes, developing accurate computable phenotypes remains a time-consuming and challenging process due to the heterogeneity, imprecision, and high dimensionality of EHR data.[1,2,6–9]

Various rule-based and machine learning (ML) approaches have been developed for generating computable phenotypes.[7] Due to the high-dimensional, messy, noisy data that constitute EHRs, many studies have developed ensemble or deep learning methods to train accurate models.[10–17] Algorithms employed in these studies (e.g. random forests and neural networks) generally can perform well in classification but are often limited in their *interpretability*, a subjective concept defined as the extent to which a model can be understood and/or its behavior interpreted by a user.[18–22]

Many have noted that interpretability is a key feature for EHR-based ML models.[23,24] For black-box ML methods, post-hoc approaches can estimate the impact of each feature.[25–28] However, it is advantageous to be able to explicitly understand *why* a computable phenotype is positive or negative for an individual patient.[29,30] Such concise models are easier to robustly incorporate within existing decision-making frameworks because clinicians can corroborate or second-guess predictions, ultimately leading to trust and facilitating an overall higher quality of clinical decision making. In addition, interpretable models may be more predictably adjusted as clinical practices change over time or models are applied to new settings. For these reasons, the FDA’s proposed regulatory framework for the evaluation of automated clinical decision support systems incorporates whether clinicians can “independently review the basis for [a model’s] recommendations” as critical to risk stratification of future ML deployments in medicine.[31] See Supplementary Materials for further background.

In this paper, we improved and then applied the feature engineering automation tool (FEAT) to generate computable phenotypes that are both accurate and interpretable.[32–34] FEAT uses a genetic programming approach for symbolic regression.[35] It learns interpretable feature representations in tandem with fitting a classification model. The representations are evolved using a population-based Pareto optimization algorithm that jointly optimizes model discrimination and complexity.[36,37] To our knowledge, this is the first work to explore the application of symbolic regression with Pareto optimization to EHR phenotyping.

We applied FEAT to EHR data targeting primary aldosteronism (PA), the most frequent cause of secondary hypertension.[38] Epidemiological studies suggest that PA affects ∼1% of US adults, but recent literature demonstrates it is under-screened for and under-diagnosed.[39–44] Using FEAT, we have developed preliminary computable phenotypes for identifying patients for whom guidelines recommend PA screening, patients with hypertension with unexplained hypokalemia (HTN-hk) or apparent treatment-resistant hypertension (aTRH).[39] PA is thought to be responsible for these phenotypes in up to 20% of these patients.[39–41,45] We expect that identifying such patients who should be screened for PA could drive improvements in their care.

## MATERIALS AND METHODS

### Benchmark Data

To benchmark changes to FEAT, we applied variant methods to 20 classification tasks in the Penn Machine Learning Benchmark (PMLB; Supplementary Tables 1 & 2).[46]

### Patients

We studied 1,200 patients receiving longitudinal primary care in the University of Pennsylvania Healthcare System (UPHS). Subjects included had (1) at least five outpatient visits in at least three separate years between 2007 and 2017, (2) at least two encounters at one of 40 primary care practice sites, and (3) were 18 years or older in 2018. A set of 1,000 random subjects from this cohort were divided into 800 for model training and 200 for model testing. One subject in the random training set was excluded because of a mid-study change in enterprise master patient index (EMPI) identifier.

A study physician (I.A.) reviewed clinical charts and classified subjects with respect to three phenotypes of increasing complexity for hypertension related to screening guidelines for PA: hypertension, HTN-hk, and aTRH. Classification was based on JNC7 Guidelines on Prevention, Detection, Evaluation, and Treatment of High Blood Pressure.[47] Unclear cases were further reviewed by an additional study physician (D.S.H. or J.C.). See Supplementary Material for further details.

Preliminary and final expert-curated heuristics for aTRH and HTN-hk (see below) were used to identify an additional 50 subjects each for model training and model testing, respectively. This yielded a total of 899 subjects for the training set and 300 subjects in the testing set. This study protocol was reviewed and approved by University of Pennsylvania Institutional Review Board (#827260).

### Clinical Data

We extracted 331 features from the EHR clinical data repository Penn Data Store and EPIC Clarity reporting database (Supplementary Tables 3 - 8). Demographic and encounter features included age, race, sex, categorized distance from zip code 19104, weight, BMI, blood pressures, and number of elevated blood pressures. Longitudinal features were aggregated as minimum, maximum, median, standard deviation, and skewness. The 34 most common laboratory test results (complete metabolic panel, complete blood count with differential, lipids, thyroid stimulating hormone, and hemoglobin A1c) with < 33% missingness were summarized as minimum, maximum, median, 1^st^ quartile, and 3^rd^ quartile. Diagnosis codes for hypertension, associated comorbidities, and other indications for anti-hypertensive medications were aggregated and summarized as median per year and sum. Medication prescriptions were summarized as the number of days prescribed for each antihypertensive class and the counts of encounters while prescribed 1, 2, 3, or 4 or more anti-hypertensive medications, summarized as sum, median, standard deviation, and skewness, as well as the sum of encounters with elevated blood pressures. Regular expressions, adapted with modifications from Teixeira et. al.,[48] were applied to clinical notes to identify mentions of ‘hypertension’ and variants thereof, summarized as counts. Features with values outside of physiologically reasonable ranges, less than 5% non-zero counts, or variance less than 0.05 were excluded. Missing values were median imputed.

### Construction of expert-curated heuristics

Next, computable phenotypes (heuristics) were manually curated for the three target phenotypes by expert review of EHR data and several iterations of proposing, applying, and evaluating the heuristics. Heuristics were initially developed from the set of random training patients. A preliminary set of heuristics for HTN-hk and aTRH were used to identify 50 patients, and iteratively evaluated and updated. Thus, final heuristics were developed from the entire set of 799 random and 100 targeted training patients. Final heuristics were then used to identify an additional 100 patients for the held-out testing set.

The heuristic designed for hypertension queried for a history of two or more diagnosis codes for hypertension (International Classification of Diseases [ICD]-9: 401.*, 405.*; ICD-10: I10.*, I15.*). For HTN-hk, we labeled patients with at least two diagnosis codes for hypokalemia (ICD-9: 276.8; ICD-10: E87.6), or at least two outpatient encounters with low blood potassium (< 3.6 mmol/L), or at least two prescriptions for an oral potassium supplement. For aTRH, we modified a previously reported heuristic[49] to label patients (1) with documentation of at least 2 out of 5 consecutive outpatient encounters with elevated blood pressure (systolic blood pressure ≥ 140 mmHg or diastolic blood pressure ≥ 90 mmHg) while on antihypertensive medications from 3 distinct classes for at least 30 days prior to the elevated blood pressures or (2) prescribed four or more antihypertensive drug classes for at least 30 days. Exclusion criteria for aTRH included patients with a diagnosis code for heart failure or transplant (ICD-9: 428.*, V42.1; ICD-10: 150.*, Z94.1) or moderate to severe chronic kidney disease (estimated glomerular filtration rate [Modification of Diet in Renal Disease; MDRD]) < 45 mL/min/1.73 m^2^) prior to meeting the above criteria.

### Feature Engineering Automation Tool (FEAT)

We adapted a recent method for learning informative feature representations called FEAT (v0.4.2) for largely automated clinical phenotyping (Figure 1; https://lacava.github.io/feat).[32–34] For this task, we are interested in learning a classification model from a set of *N* paired samples, {(*y*_*i*_, ***x***_*i*_), i = 1, …, N}, with binary labels *y* ∈ {0,1} and attributes *x* ∈ ***R***^”^. FEAT attempts to learn a set of features for a logistic regression model of the form

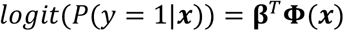

where **ϕ**(***x***) is a *p*-dimensional vector of transformations of ***x*** learned from FEAT’s optimization process. The coefficients β = [β_1_, …, **β**_*p*_] are associated with each of these transformed features.

**Figure 1.**
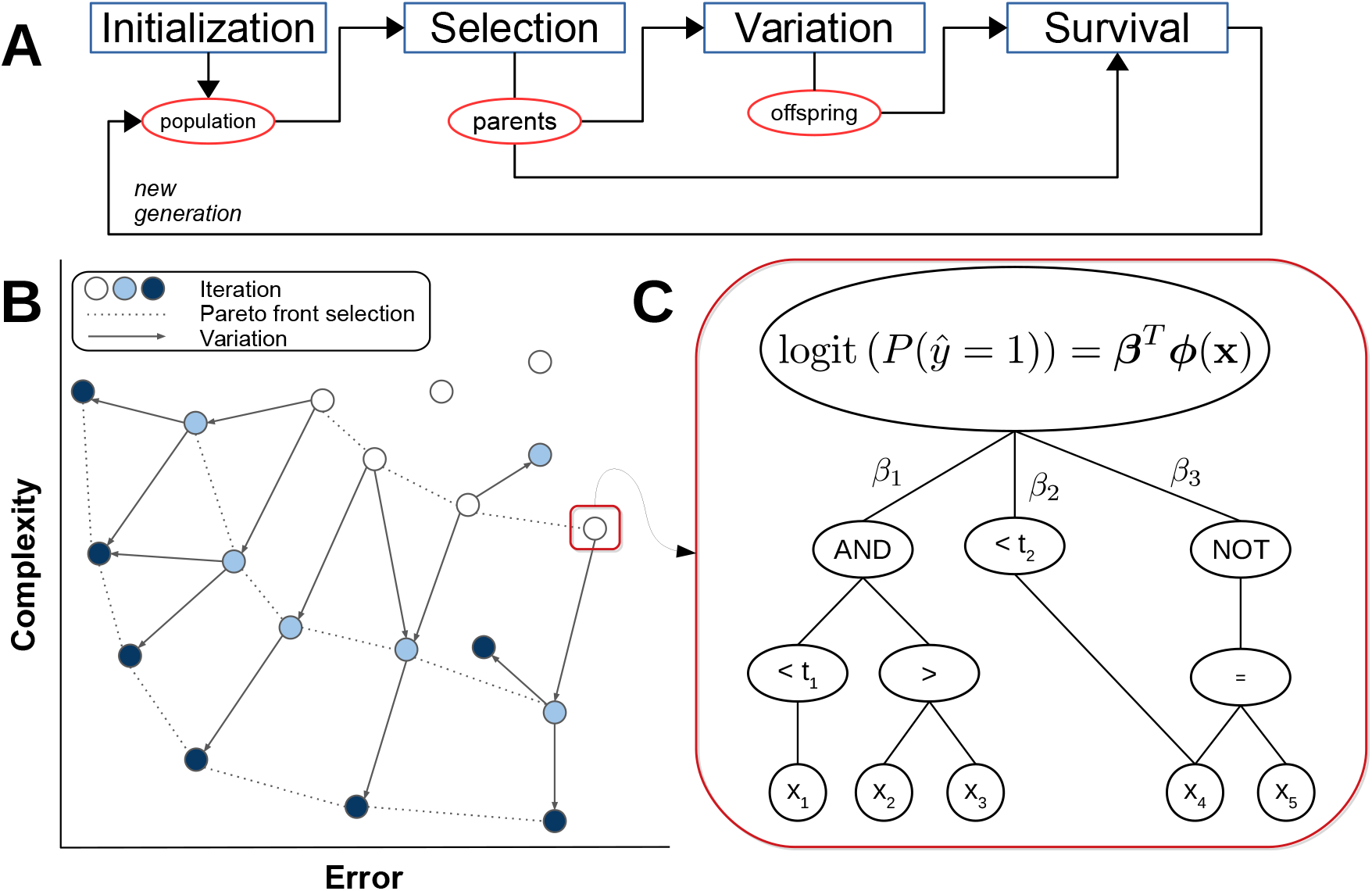
How FEAT works. (A) Steps in the genetic programming process. Candidate models are initialized in a population; the best models (parents) are selected via epsilon-lexicase selection; offspring are created by applying variation operations to the parents; and then parents and offspring compete in a survival step using NSGA-II [22]. The process then repeats. (B) The evaluation of a candidate models’ complexity and performance in Pareto Optimization framework in the Survival step. (C) Example model in which input features are transformed by logical functions with or without threshold operators.

For the purposes of learning interpretable models, we restricted the transformation operators to Boolean functions: <, >, AND, OR, NOT. This limits the search space to representations consisting of these operators and the input features. For inequalities, we included operators that use Gini impurity to choose the split threshold for each feature in an equivalent way to classification trees. Note that because the optimization process includes mutation to or insertion of new input features, it allows for non-greedy search to occur to find the best fit for the problem at hand, in contrast to decision trees.

To encourage model parsimony, we modified FEAT in two distinct ways. First, to handle high-dimensional data, rather than fitting a multivariate linear model to all the data at the start of optimization, we sampled the input data based on univariate logistic regression coefficients. Second, we added a post-run simplification procedure to shrink the final feature representation without significantly altering its behavior. This post-run simplification procedure consists of 1) explicitly removing redundant serial logical operators, 2) adaptively pruning highly correlated components of representations, and 3) applying random deletion mutations to the features in a hill-climbing fashion. See the Supplementary Methods for further details.

### Comparator Methods

To assess how FEAT models compare to conventional ML models, we applied five supervised classifiers: LASSO-penalized logistic regression (LR L1), ridge-penalized logistic regression (LR L2), decision tree (DT), random forest (RF), and Gaussian Naïve Bayes (GNB). Hyperparameters for each of the models were optimized using 5-fold nested cross-validation. All of the comparator methods were implemented using Scikit-learn.[50] We report the mean test area under the precision-recall curve (AUPRC) and area under the receiver-operating curve (AUROC) for all experiments. AUPRC is calculated as average precision (see *sklearn*.*metrics*.*average_precision_score*, scikit-learn version 0.23.2). We also compared the size of the final models, defined for tree-based methods (FEAT, decision tree, and random forest) as the total number of nodes in the trees and defined for the linear methods and GNB as the number of predictors with non-zero coefficients. Models’ performance and size were compared using paired Wilcoxon rank-sum tests. Model thresholds were selected in the training set to achieve a positive-predictive value (PPV) in the longitudinal, primary care cohort of 0.70. Study code, including full environment specification, is available in the repository https://bitbucket.org/hermanlab/ehr_feat/.

## RESULTS

### Development of automated phenotyping method

To automatically construct computable phenotypes whose outputs are interpretable by clinicians, we extended FEAT to better implement Boolean logic, added procedures to encourage model parsimony, and developed approaches for improving training robustness. To evaluate these modifications, we applied them to benchmark datasets[46] that were similar in shape to our EHR data. We found that restricting operators and simplifying models did not significantly impair classification performance but substantially decreased the size of resulting models (Supplementary Fig. 1; p = 7.2×10^−9^). Detailed results are available in the Supplementary Material.

### Learning expert-curated computable phenotypes

We next applied our optimized FEAT method to a training set of 899 subjects to learn to recapitulate the expert-curated heuristics for hypertension, HTN-hk, and aTRH. We evaluated each heuristic in 50 trials of 5-fold cross-validation on shuffled training datasets and averaged test scores across folds (Figure 2, top row; Table 1). Across all three heuristics, FEAT models achieved higher AUPRC (*p* < 0.001; Supplementary Fig. 2) than all other models except RF. FEAT models were smaller than all other models (*p* < 1×10^−6^) except decision trees.

**Table 1.** Computable phenotype discrimination and size for each target phenotype in training cross-validation.

| Phenotype | Method | Median CV<br>AUPRC (IQR) | Median CV<br>AUROC (IQR) | Median Size (IQR) |
| --- | --- | --- | --- | --- |
| HTN Heuristic | GNB | 0.93 (0.01) | 0.96 (0.00) | 331.00 (0.00) |
|  | DT | 0.99 (0.00) | 1.00 (0.00) | 4.60 (0.00) |
|  | LR L1 | 1.00 (0.00) | 1.00 (0.00) | 52.30 (5.90) |
|  | LR L2 | 0.99 (0.00) | 0.99 (0.00) | 330.80 (0.00) |
|  | RF | 1.00 (0.00) | 1.00 (0.00) | 8760.40 (6278.30) |
|  | FEAT | 1.00 (0.00) | 1.00 (0.00) | 8.20 (0.95) |
| HTN-Hypokalemia<br>Heuristic | GNB | 0.47 (0.02) | 0.86 (0.00) | 331.00 (0.00) |
|  | DT | 0.95 (0.02) | 0.99 (0.01) | 10.80 (1.60) |
|  | LR L1 | 0.98 (0.01) | 1.00 (0.00) | 65.10 (18.05) |
|  | LR L2 | 0.87 (0.02) | 0.96 (0.01) | 330.80 (0.00) |
|  | RF | 1.00 (0.00) | 1.00 (0.00) | 1855.80 (1640.80) |
|  | FEAT | 0.99 (0.01) | 1.00 (0.00) | 18.30 (2.55) |
| Resistant HTN<br>Heuristic | GNB | 0.58 (0.02) | 0.92 (0.01) | 331.00 (0.00) |
|  | DT | 0.67 (0.05) | 0.89 (0.03) | 27.00 (3.20) |
|  | LR L1 | 0.84 (0.02) | 0.95 (0.01) | 70.50 (16.90) |
|  | LR L2 | 0.81 (0.02) | 0.94 (0.01) | 330.80 (0.00) |
|  | RF | 0.91 (0.02) | 0.99 (0.00) | 27235.00 (13392.00) |
|  | FEAT | 0.91 (0.02) | 0.98 (0.01) | 14.20 (1.70) |
| HTN Diagnosis | GNB | 0.93 (0.00) | 0.95 (0.00) | 331.00 (0.00) |
|  | DT | 0.91 (0.01) | 0.94 (0.01) | 51.00 (5.20) |
|  | LR L1 | 0.98 (0.00) | 0.98 (0.00) | 23.20 (5.50) |
|  | LR L2 | 0.97 (0.00) | 0.98 (0.00) | 330.80 (0.00) |
|  | RF | 0.98 (0.00) | 0.99 (0.00) | 20397.60 (19474.10) |
|  | FEAT | 0.98 (0.00) | 0.98 (0.00) | 13.90 (2.90) |
| HTN-Hypokalemia<br>Diagnosis | GNB | 0.38 (0.01) | 0.86 (0.02) | 331.00 (0.00) |
|  | DT | 0.63 (0.06) | 0.88 (0.04) | 30.20 (2.40) |
|  | LR L1 | 0.80 (0.02) | 0.94 (0.02) | 68.10 (15.30) |
|  | LR L2 | 0.73 (0.02) | 0.91 (0.02) | 330.80 (0.00) |
|  | RF | 0.84 (0.02) | 0.98 (0.01) | 13443.40 (8241.30) |
|  | FEAT | 0.82 (0.03) | 0.97 (0.01) | 16.20 (2.70) |
| Resistant HTN<br>Diagnosis | GNB | 0.46 (0.02) | 0.90 (0.02) | 331.00 (0.00) |
|  | DT | 0.41 (0.06) | 0.78 (0.04) | 56.80 (7.40) |
|  | LR L1 | 0.69 (0.04) | 0.93 (0.02) | 52.90 (17.95) |
|  | LR L2 | 0.69 (0.04) | 0.91 (0.01) | 330.80 (0.00) |
|  | RF | 0.75 (0.02) | 0.96 (0.00) | 38835.00 (20521.10) |
|  | FEAT | 0.69 (0.05) | 0.94 (0.01) | 9.80 (1.80) |

**Figure 2.**
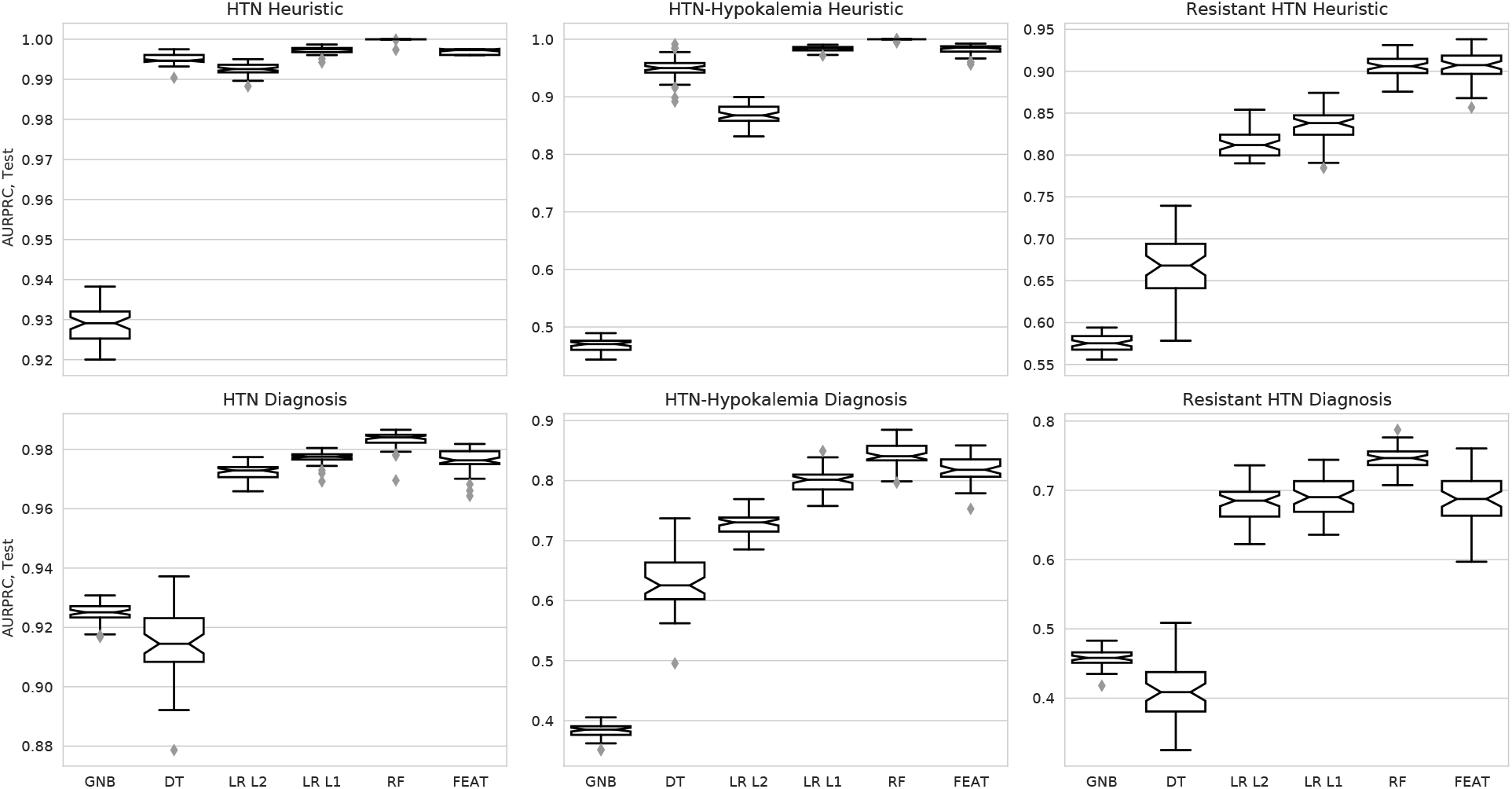
Estimating model discrimination by cross-validation. AUPRC scores for phenotyping models trained in 5-fold cross-validation over 50 iterations, each averaged across testing folds. Each subplot represents a different training outcome; heuristics are shown in the top row, and chart-review diagnoses are shown in the bottom row.

### Automated learning of computable phenotypes

Next, we compared the performance of models trained to predict the chart-review phenotypes (Figure 2, bottom; Table 1), which were present in 423 (47%), 93 (10%), and 103 (11%) subjects, respectively. Across all phenotypes, FEAT models achieved AUPRC scores that were higher than GNB, LR L2, and DT models (*p* < 0.001; Supplementary Fig. 2), comparable to LR L1 models (*p* > 0.99), and slightly lower than RF models (*p* < 1e-6). These relationships were consistent across phenotypes, except that FEAT models appeared to also outperform LR L1 for HTN-hk. FEAT models were smaller than all other models including decision tree models (*p* < 1e-6); models were on average approximately 1800 times smaller than RF models and 2.9 times smaller than LR L1 models. We next explored the trade-off between model performance and complexity for heuristic and chart-review trained models (Figure 3). The FEAT models clustered near the high-performance, low-complexity region (top left) of this tradeoff space, indicating that they achieved a relatively efficient trade-off between these two objectives.

**Figure 3.**
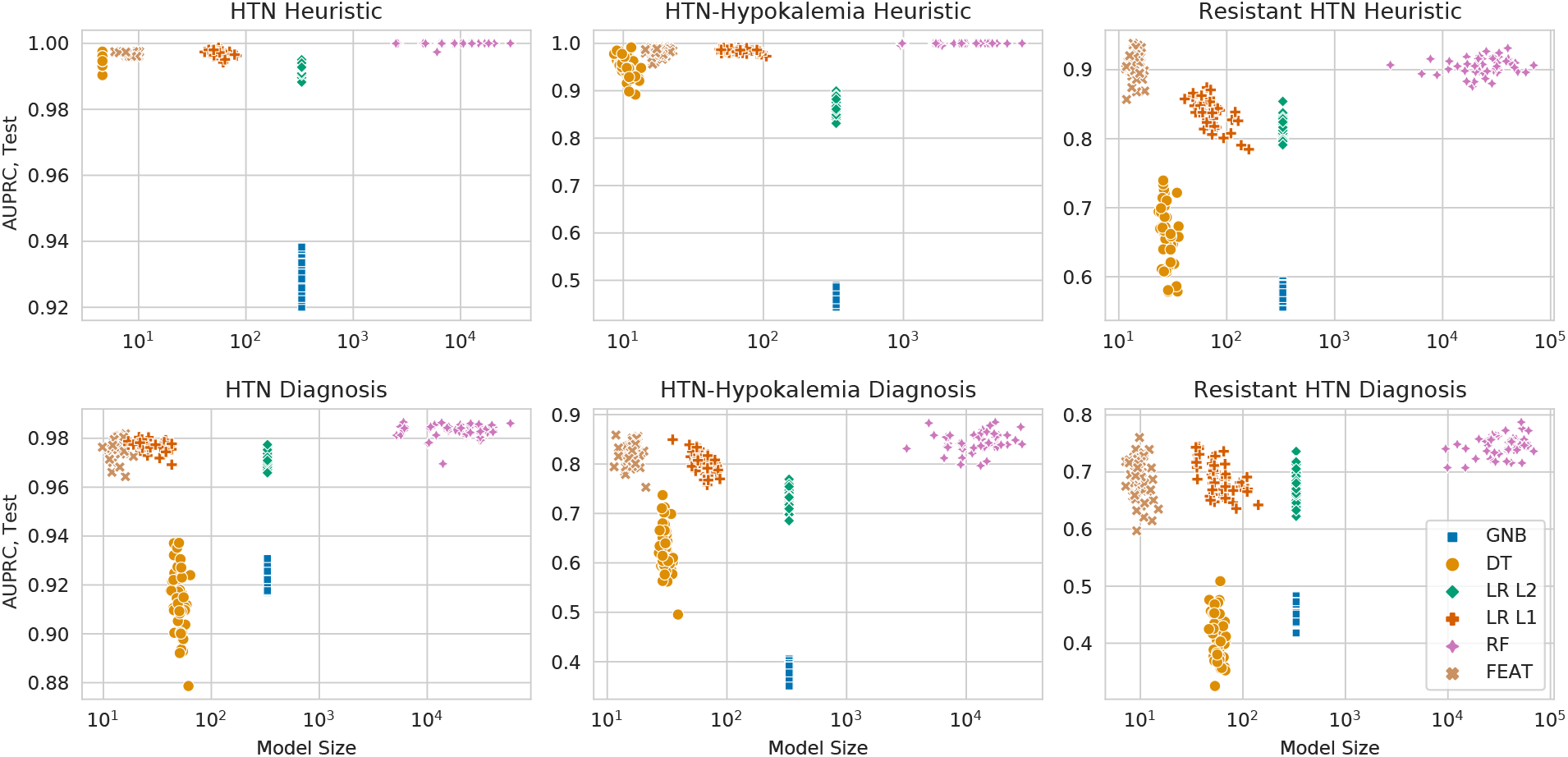
The tradeoff between model discrimination and complexity. Each point shows the cross-validation testing AUPRC (y-axis) and size (x-axis) for models trained in 50 repeat trials for each method. Each subplot represents a different expert-curated heuristic (top row) or chart review phenotype (bottom). The ideal model is discriminative and simple, meaning it is near the top left corner.

For the most complex phenotype, aTRH, FEAT models achieved a median AUPRC of 0.69 (interquartile range [IQR]: 0.05) with a median size of 9.8 (IQR: 1.8). These models showed reasonable discrimination across all potential decision thresholds, as depicted by PRC and ROC (Figure 4). Of note, the expert-curated heuristic demonstrated superior discrimination to all ML models at its single operating point.

**Figure 4.**
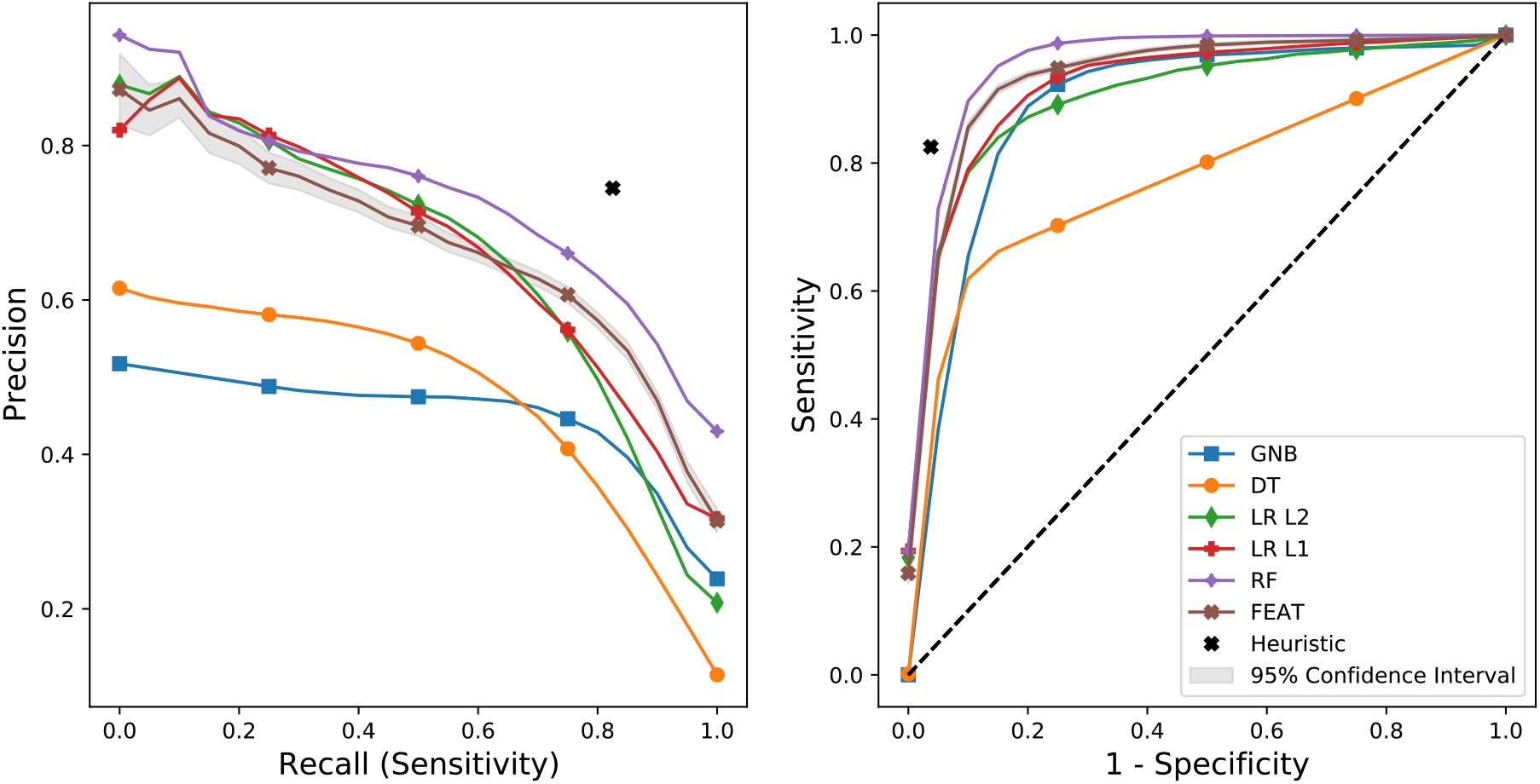
Model precision-recall and receiver-operating curves. Precision-recall curves (left) and receiver-operating curves (right) for phenotyping models trained to predict chart review classifications for aTRH. Values shown are means of test performance in 5-fold cross-validation iterated 50 times.

### Assessment of model generalization and clinical utility

Next, we applied the methods refined by cross-validation to learn models from the entire training set and assessed their performance on a held-out test set of 300 subjects, including 185 (61%), 79 (26%), and 73 (24%) subjects for each chart-review phenotype. Model performance and size (Table 2) were consistent with cross-validation estimates. Most appeared to have slightly better AUPRC than in cross-validation, likely due in part to the higher enrichment for cases in the testing cohort. For chart-reviewed hypertension, HTN-hk, and aTRH, the FEAT models demonstrated AUPRC scores of 0.99, 0.96, and 0.80, and AUROC scores of 0.99, 0.98, and 0.94, respectively. As compared to the expert curated heuristics, the FEAT models’ AUPRC was 0.15 (18%) higher for HTN-hk and within 0.02 (2%) for hypertension and aTRH.

**Table 2:** Final model discrimination in test set and size.

| Phenotype | Method | Test AUPRC | Test AUROC | Size |
| --- | --- | --- | --- | --- |
| HTN Heuristic | GNB | 0.94 | 0.95 | 331 |
|  | DT | 1.00 | 1.00 | 5 |
|  | LR L1 | 1.00 | 1.00 | 56 |
|  | LR L2 | 1.00 | 0.99 | 331 |
|  | RF | 1.00 | 1.00 | 3106 |
|  | FEAT | 1.00 | 1.00 | 12 |
| HTN-Hypokalemia Heuristic | GNB | 0.73 | 0.85 | 331 |
|  | DT | 1.00 | 1.00 | 13 |
|  | LR L1 | 1.00 | 1.00 | 76 |
|  | LR L2 | 0.98 | 0.98 | 331 |
|  | RF | 1.00 | 1.00 | 1106 |
|  | FEAT | 1.00 | 1.00 | 24 |
| Resistant HTN Heuristic | GNB | 0.66 | 0.89 | 331 |
|  | DT | 0.82 | 0.92 | 27 |
|  | LR L1 | 0.92 | 0.96 | 101 |
|  | LR L2 | 0.93 | 0.97 | 331 |
|  | RF | 0.96 | 0.99 | 2648 |
|  | FEAT | 0.94 | 0.98 | 16 |
| HTN Diagnosis | GNB | 0.96 | 0.96 | 331 |
|  | DT | 0.97 | 0.97 | 43 |
|  | LR L1 | 1.00 | 0.99 | 32 |
|  | LR L2 | 0.99 | 0.98 | 331 |
|  | RF | 1.00 | 0.99 | 67276 |
|  | FEAT | 0.99 | 0.99 | 18 |
|  | Expert Heuristic | 0.97 | 0.97 | - |
| HTN-Hypokalemia Diagnosis | GNB | 0.60 | 0.81 | 331 |
|  | DT | 0.75 | 0.86 | 33 |
|  | LR L1 | 0.95 | 0.98 | 29 |
|  | LR L2 | 0.92 | 0.96 | 331 |
|  | RF | 0.96 | 0.99 | 16256 |
|  | FEAT | 0.96 | 0.98 | 8 |
|  | Expert Heuristic | 0.81 | 0.96 | - |
| Resistant HTN Diagnosis | GNB | 0.57 | 0.86 | 331 |
|  | DT | 0.23 | 0.47 | 67 |
|  | LR L1 | 0.74 | 0.87 | 130 |
|  | LR L2 | 0.78 | 0.91 | 331 |
|  | RF | 0.89 | 0.96 | 119760 |
|  | FEAT | 0.80 | 0.94 | 11 |
|  | Expert Heuristic | 0.82 | 0.96 | - |

To further evaluate the utility of the resulting models, we selected diagnostic interpretive thresholds. For the goal of identifying patients that should be screened for PA using models predicting aTRH, we targeted a model PPV ≥ 0.70 amongst primary care patients. Assuming that 20% of aTRH patients have PA, we expect that approximately 1 in 7 aTRH model-positive patients would have PA. We also assumed an aTRH prevalence of 7.5%, based on the frequency observed in our training set and meta-analyses.[51] This resulted in the selection of a threshold of 0.40, which corresponded to a sensitivity of 0.82 in training. Among the 200 randomly drawn test subjects, this FEAT model yielded an adjusted PPV of 0.70 and sensitivity of 0.62. In comparison, the heuristic showed an adjusted PPV of 0.87 and sensitivity of 0.92. To evaluate FEAT on a richer set of cases, we also assessed its performance on 100 test patients selected by the final aTRH or HTN-hk heuristics. In this set, the final FEAT model had a PPV of 0.79 and the expert heuristic a PPV of 0.83.

### Model interpretability

Finally, we evaluated the relative interpretability of the resulting models, focusing on the models for predicting aTRH. The final FEAT model was concise and interpretable (Figure 5). The FEAT model assigned risk according to the following factors, in order of absolute coefficient magnitudes: first, a history of more than one encounter while prescribed three or more anti-hypertensive medications (β = 1.33); second, a mean systolic blood pressure above 128.6 mmHg (β = 0.95); third, a history of low variability (standard deviation) in the number of encounters while prescribed two anti-hypertensive medications each year (β = −0.52); fourth, a history of a median of 1.25 or more encounters per year while prescribed four or more hypertension medications (β = 0.49); fifth, more than 40 mentions of hypertension in patient notes (β = 0.42); and sixth, a maximum total calcium greater than 10.1 mg/dL (β = 0.40). To investigate the factors underlying the maximum calcium feature, we explored its associations.

**Figure 5.**
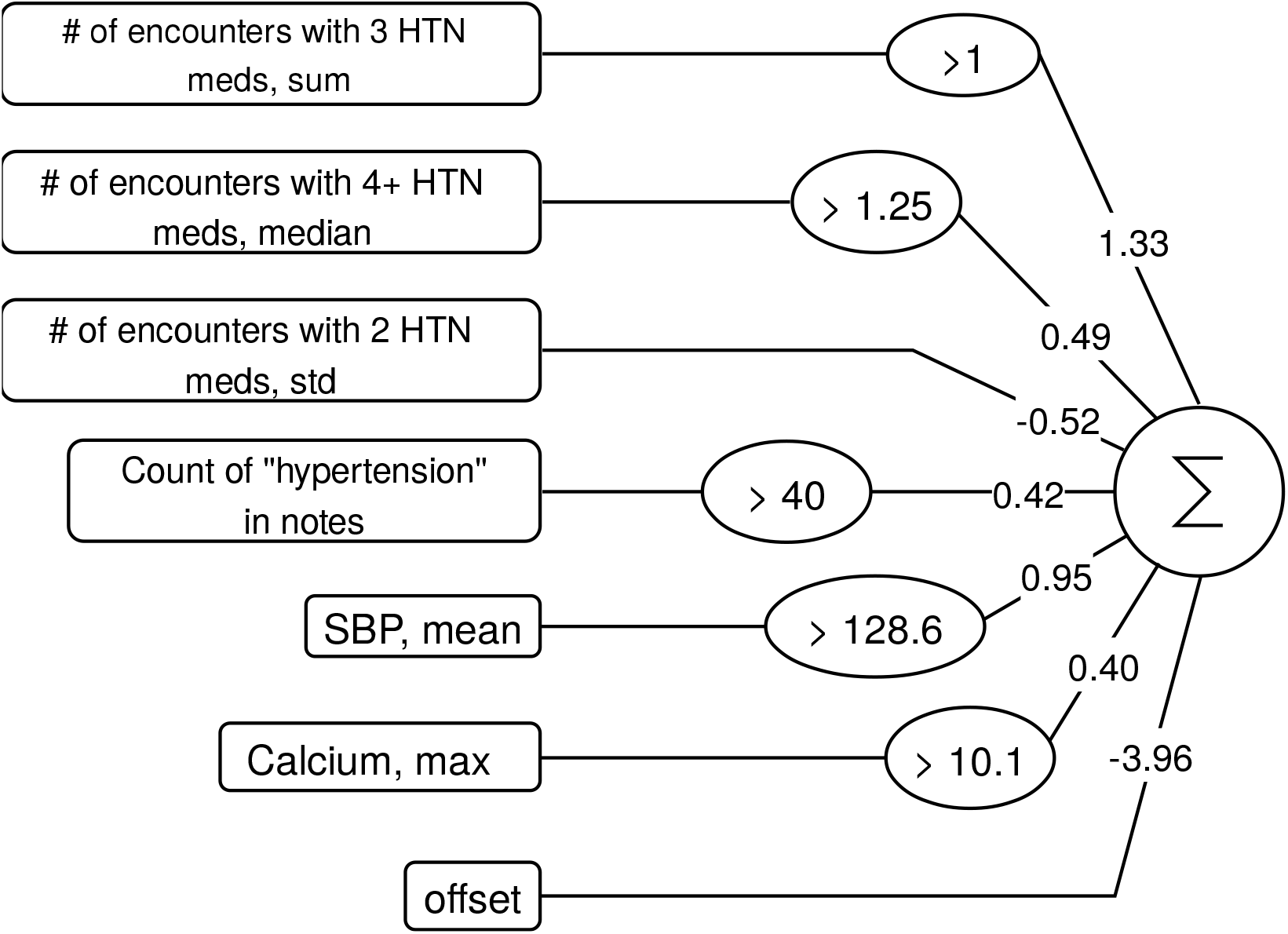
FEAT model trained to predict apparent treatment-resistant hypertension. The input features are shown on the left followed by the learned operations, the multiplication coefficients, and the summation. Note, the subsequent logit transformation and interpretive threshold is not depicted.

We found that subjects with aTRH were in fact more likely to have an elevated maximum calcium (OR=4.4, p=4×10^−9^) and that these elevations were in turn associated with the number of days prescribed thiazide diuretics (OR=1.5 per SD, p=3×10^−6^) or beta-blockers (OR=1.4, p=2×10^−4^).

None of the other derived models can be described in such compact, clear language. So, to compare and contrast FEAT with other methods, we calculated SHAP values[28] for the test subjects. SHAP values summarize the impact of input variables on model outputs by generating an additive feature attribution model. Positive and negative SHAP values indicate a marginal increase and decrease in predictions, respectively. The summary plots for SHAP values (Figure 6A,C) depict the distribution of SHAP values relative to the magnitude of each input variable, with each dot representing a single test subject. The decision plots of SHAP values (Figure 6B,D) illustrate how each feature contributes to predictions for individual subjects.

**Figure 6.**
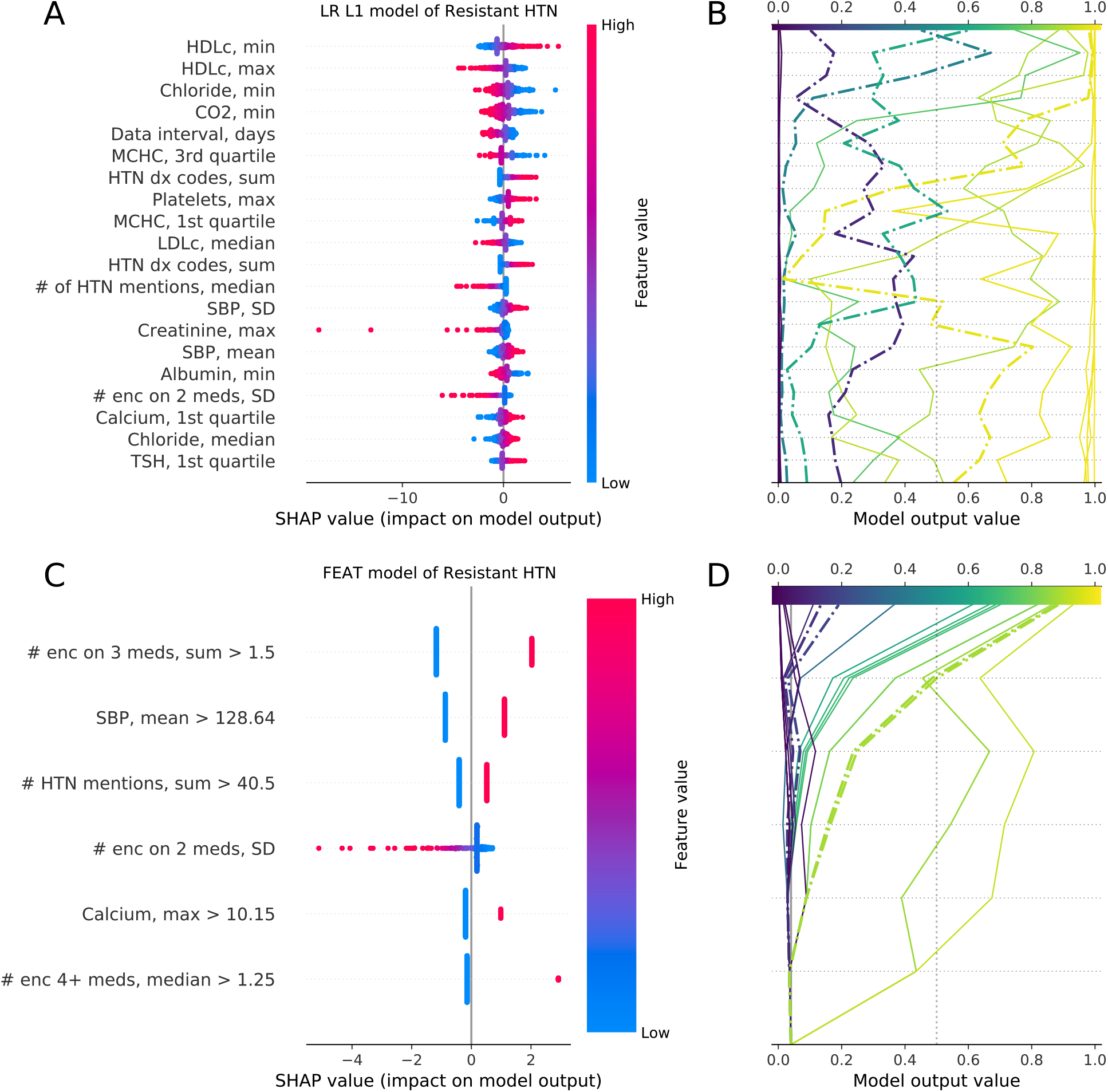
SHAP plots for explaining models. SHAP summary (A) and decision (B) plots for the LR L1 and summary (C) and decision (D) plots for the FEAT models. The summary plots (A,C) describe the most important features, ranked by the mean absolute SHAP value in the test data. Each point represents a subject; its color reflects the relative feature value and the location along x-axis its impact on the subject’s model output. The lines in the decision plots (B,D) show model predictions for a sample of 10 positive and 10 negative predictions, with dash-dotted lines indicating misclassifications. The summary and decision plots are aligned vertically, such that the feature labels in the summary plots correspond to the incremental changes in the adjacent decision plot lines, indicating the feature responsible for the change in the model score at each level.

The FEAT summary plot (Figure 6C) reflects the simplicity of the FEAT model. For the five dichotomized features, each patient’s prediction is either increased or decreased by a fixed increment. The one continuous feature affects each patient distinctly, but has a clear directionality, i.e. high variability in the number of encounters on two anti-hypertensive medications decreases the prediction. These simple effects translate into intuitive interpretations for individual subjects as to *why* the model is calling them positive or negative (Figure 6D). The positive-slope increases in model output show that most patients predicted to be positive have had multiple encounters while prescribed three anti-hypertensive medications. They also either have elevated mean systolic blood pressure and many mentions of hypertension in notes or multiple encounters per year while prescribed four or more anti-hypertensive medications.

In contrast, the LR L1 (Figure 6A,B) and RF (Supplementary Fig. 3) summary and decision plots reflect much more complicated models, in which many features contribute to the prediction scores. The summary plots show the modest effect of each of the 20 displayed features, which demonstrated the highest model coefficients or variable importance. The decision plots demonstrate that each patient has a distinct *reason* for a positive or negative prediction, determined by a combination of many features. In addition, there is also considerable signal from the features not depicted, as evident in the variable, non-zero intercepts between each patient’s line and the model output value x-axis. Notably, for the LR L1 model many of the top features (e.g. minimum HDL cholesterol) are not intuitively linked to the phenotype, likely due to feature co-linearity. To address this, we also calculated LR L1 SHAP values after adjusting for feature covariance (Supplementary Fig. 4A,B). After adjustment, the top features (e.g. # enc 4+ meds, median) more closely matched clinical intuition. However, the relationships between features and SHAP values remained complex, including a large number of features with small individual effects. For the sake of comparison, we also accounted for co-linearity in the FEAT model (Supplementary Fig. 4C,D). These FEAT model explanations remained similarly concise.

Of note, FEAT’s emphasis on small model size does have costs. For instance, some patients with heart failure or chronic kidney disease were misclassified as positive for aTRH by the FEAT model (Figure 6D). In contrast, the LR L1 model lowers prediction scores based on maximum creatinine or heart failure diagnosis codes (Supplementary Fig. 4A). Such features were commonly observed in FEAT models along the Pareto-optimal front during training, but these models were ultimately not selected because of their higher overall complexity.

## DISCUSSION

We developed a method to automate the construction of EHR computable phenotypes and applied this method to find patients that should be screened for PA. Conventional approaches for manually building computable phenotypes cannot scale to the expanse of clinical use cases.

However, it may be possible to automate their construction by embedding the design goals for such heuristics into ML approaches. In the manual training of computable phenotypes, experts incorporate clinical knowledge to develop intuitive sets of rules. Our goal in developing and applying FEAT is to automate this process by generating symbolic models that are both highly accurate and interpretable by clinicians.

We compared FEAT’s ability to learn computable phenotypes to that of expert heuristic curation and conventional ML approaches. The models FEAT constructed were more concise and interpretable than those of other ML approaches that achieved similar discrimination. In fact, the FEAT models matched the discriminative performance of other models across various tasks, except for the RF model for the most complicated phenotype, aTRH. In this case, the FEAT model showed slightly lower discrimination than the RF model but was much more interpretable.

In comparison to expert-curated heuristics, the FEAT models showed better discrimination for hypertension and HTN-hk. Notably, FEAT’s model for HTN-hk performed on par with other ML models and better than the heuristic while only consisting of 8 components. However, FEAT models showed lower discrimination for aTRH. This underperformance was expected for several reasons. First, the comparison between FEAT and the expert heuristic was biased because the heuristic was constructed and refined using the entire training set and was used to identify many of the affected test subjects, likely inflating its observed performance in training cross-validation and testing. Second, the FEAT method was not empowered to learn the temporal relationships between features that enabled the expert heuristic to achieve high specificity, such as the minimum time interval between meeting hypertensive medication criteria and assessment for persistently elevated blood pressure. We expect that future improvements to this feature representation learning method may enable native identification of such temporal relationships from longitudinal EHR data.

The model that FEAT learned to identify patients with aTRH was both accurate and understandable. Its components largely matched those of the expert heuristic and were consistent with clinical intuition. FEAT learned to combine complementary sources of information, including medication, vitals, laboratory results, and concepts from notes. Finally, it learned an unexpected but clinically intuitive and valuable rule related to maximum blood calcium levels. Anti-hypertensive medications, particularly diuretics, can dysregulate calcium homeostasis. In addition, hyperparathyroidism, which causes elevated blood calcium, is associated with hypertension. We suspect this rule enabled the model to identify a few affected subjects, on intensive anti-hypertensive regimens and/or with underlying hyperparathyroidism, who were missed by the conventional heuristics that considered only medication prescriptions and blood pressure.

There are several possible directions for further improving FEAT. For one, the ability of FEAT to recapitulate expert-curated heuristics suggests that simpler expert heuristics, such as anchor variables,[52] may be leveraged as teachers in a semi-supervised approach. One limitation of this work is the non-trivial, manual feature engineering upstream of FEAT. Future work could reduce this manual feature engineering by enabling FEAT to directly incorporate longitudinal data. Although application to raw input features would increase the search space considerably, this would enable learning of temporal relationships essential for prediction of phenotypes like aTRH. Finally, this approach could benefit considerably from learning on top of standard frameworks for representing and querying both clinical data and expert clinical knowledge.[53,54] The incorporation of expert knowledge would improve search efficiency while maintaining interpretability.

## CONCLUSION

In summary, FEAT can effectively learn highly accurate and interpretable computable phenotypes. We expect that this approach will ultimately empower experts to much more efficiently construct computable phenotypes, facilitating widespread implementation of computable phenotype-triggered clinical decision support and translational research.

## Data Availability

Public data used for benchmarking FEAT is available from www.github.com/EpistasisLab/pmlb. Clinical data underlying this article cannot be shared publicly to protect the privacy of the subjects. Upon request and subject to appropriate approvals, it will be shared by the corresponding author.

## ACKNOWLEDGEMENTS

We would like to thank Debbie Cohen for helpful discussions about secondary hypertension.

## AUTHOR CONTRIBUTIONS

WL, PCL, and DSH designed the study and wrote the manuscript. WL, PCL, JHM, and DSH developed the method. XD, PS, and DSH collected, wrangled, and transformed the clinical data. IA, JBC, and DSH performed the chart review. WL and PCL performed the analyses. All authors approved the final manuscript.

## DATA AVAILABILITY

The data underlying this article cannot be shared publicly to protect the privacy of the subjects. Upon request and subject to appropriate approvals, it will be shared by the corresponding author.

