## Supplementary Material for "Application of concise machine learning to construct accurate and interpretable EHR computable phenotypes"

### INTERPRETING PREDICTION MODELS

There are two overarching approaches to interpretable modeling. The first is to apply a post-hoc analysis tool to a black box model that determines which factors are relevant to the model's predictions.[1] Examples of post-hoc methods include permutation importance[2,3], LIME[4], and SHAP[5]. SHAP values in particular can be very useful for describing how a black-box model behaves under specific input conditions.[6] However, these approaches do not describe the *mechanism* by which factors result in the predictions. Furthermore, since these tools cannot describe the behavior of the model over all input conditions, it is challenging to predict model behavior as inputs change.[7]

The second approach to interpretable modeling is to focus on learning concise models that are self-explanatory. As Lundberg et al. put it, "the best explanation of a simple model is the model itself."[5] The most commonly used method in this category is logistic regression, often employed with regularization approaches, such as the least absolute shrinkage and selection operator (LASSO) and ridge regression.[8,9] Decision trees and Bayesian rule lists can generate interpretable models when constrained to small tree depths and low rule count, respectively. Yet these approaches are limited in that smaller models may not adequately represent complex data relationships and larger models are not practically interpretable.[10] In regularized regression and pruned decision trees, the trade-off between simplicity and explanatory power is left to be tuned by the user. More sophisticated strategies can characterize the trade-off between model complexity and model accuracy, such as Pareto optimization with symbolic regression.[11] *Symbolic regression* is a method of learning the functional form and parameters of a model using a randomized, heuristic search process such as evolutionary computation.[12] *Pareto optimization* refers to a multi-objective optimization process in which preference relations between models are determined by their closeness to the "Pareto front", which is a set of points that represent the best observed trade-offs between objectives. Symbolic regression with Pareto optimization has been used to develop simple models in other domains, such as physics,[13] biology,[14] engineering.[15] To our knowledge, this is the first work to explore the application of symbolic regression with Pareto optimization to EHR phenotyping.

### EXTENDED MATERIALS AND METHODS

In this section, we detail the methodological changes made to FEAT in order to promote conciseness in the models it generates. We also describe a benchmark comparison of FEAT variants used to validate the proposed changes.

To encourage model parsimony, we modified FEAT to explicitly simplify serial logical operators, prune highly correlated feature branches, adaptively prune components of representations, and sample features based on univariate logistic regression coefficients. In this section, we give detailed descriptions of these implementations.

#### Initial feature weighting

The original FEAT algorithm initialized weights of input features according to the magnitude of their coefficient in a multivariate linear model.[16] In addition, the initial population was seeded with the multivariate linear model that was generated. Since we are interested in learning a low dimensional representation of high-dimensional data to enable interpretation, this approach was not suitable. Instead, we modified FEAT to specify initial weights of input features according to the magnitude of each feature's coefficient in a univariate logistic regression model. The initial population of linear models was constructed by sampling features according to these magnitudes and fitting a low-dimensional multivariate model.

#### Correlation Deletion Mutation

In previous work, operators for variation were introduced to make use of information about the features encoded by the representations.[17] Here, we propose an operator designed to prune representations by removing the most redundant feature. Algorithm 1 describes the process. In short, it consists of computing pairwise correlations of each feature, and among the pair that is most correlated, deleting the feature that is less correlated with the outcome variable. Algorithm 1 is used as a component of post-run simplification, described next.

Algorithm 1: Correlation Deletion Mutation

```
CorrelationDeletionMutation(ŷ(Φ(x))):
    for ϕ_i, ϕ_j in Φ(x), i ≠ j:
            corr = R^2(ϕ_i, ϕ_j)
            max_r2 = 0.0
            if corr > max_r2:
                    max_r2 = corr
                    f1 = i
                    f2 = j
    corr_f1 = R^2(ϕ_i, y)
    corr_f2 = R^2(ϕ_j, y)
```

Remove $\boldsymbol{\phi}$ from $\boldsymbol{\Phi}(\boldsymbol{x})$ with lower corr with $\boldsymbol{y}$

RETURN ($\hat{\boldsymbol{y}}_{\boldsymbol{new}}(\boldsymbol{\Phi}(\boldsymbol{x}))$, max_r2)

### Post-run Simplification

Genetic programming suffers from a phenomenon known as *bloat*, in which final equations that are produced tend to be larger than necessary for capturing their semantics.[18] Many methods exist to combat bloat,[19,20] including various pruning mutations such as Algorithm 1. A simple but effective way to reduce bloat is post-run simplification,[21] in which simplification operations are applied to the final model in a hill climbing manner. In order to avoid over-fitting, changes are only accepted if their cumulative effect on the model output is on average within a user-specified tolerance.

We introduced an automated method for simplifying final representations produced by FEAT that includes three steps. First, redundant operations, such as NOT(NOT(.)), are removed. Second, correlation deletion mutation is applied iteratively. Finally, a uniform subtree deletion operator is applied iteratively. Each iteration succeeds only if the impact on the final model is minimal, or, in the case of correlation deletions, if the features were perfectly correlated. Post-run simplification is shown concretely in Algorithm 2.

Algorithm 2: Post-run Simplification

PostRunSimplification( $\hat{\boldsymbol{y}}(\boldsymbol{\Phi}(\boldsymbol{x}))$, tol):

  $\hat{\boldsymbol{y}}(\boldsymbol{\Phi}(\boldsymbol{x}))$ – final model

  tol – tolerance for changes to output

  $\hat{\boldsymbol{y}}_{\boldsymbol{new}}$ = RemoveRedundantOperators($\hat{\boldsymbol{y}}$)

  for $|\boldsymbol{\Phi}(\boldsymbol{x})|$ iterations i:

    $\hat{\boldsymbol{y}}_{\boldsymbol{tmp}}$, max_r2 = CorrelationDeletionMutation($\hat{\boldsymbol{y}}_{\boldsymbol{new}}$)

    if ($\|\hat{\boldsymbol{y}}_{\boldsymbol{tmp}} - \hat{\boldsymbol{y}}\| / \|\hat{\boldsymbol{y}}\|$ < tol OR max_r2 == $\mathbf{1}$):

$\hat{y}_{new} = \hat{y}_{tmp}$<br>
`else: break`<br>
`for 1000 iterations:`<br>
$\hat{y}_{tmp}$`= SubtreeDeletionMutation(`$\hat{y}_{new}$`)`<br>
`if (`$\|\hat{y}_{tmp} - \hat{y}\| / \|\hat{y}\|$` < tol):`<br>
$\hat{y}_{new} = \hat{y}_{tmp}$<br>
`RETURN `$\hat{y}_{new}(\boldsymbol{\Phi}(\boldsymbol{x}))$

**Model Selection**

Due to its nature as a population-based method, FEAT's optimization process produces several candidate final models along the Pareto-optimal front. In order to choose a single final model, models are trained on 80% of available training samples and 20% of training samples are held-out for internal model validation. Then from the population of models along the Pareto front, the model with the lowest balanced log-loss in the held-out 20% of samples is selected as the final model. Due to its nature as a probabilistic algorithm, FEAT is sensitive to the random seed used in training. In order to encourage a robust final model was selected, we designed a heuristic procedure. FEAT was rerun 10 times in training, thereby yielding 10 models. Of these final models, we excluded those in the lowest quartile of validation AUPRC and then chose the smallest model. In our preliminary cross-validation analyses, we found this to result in relatively stable, discriminative, and interpretable models over 50 realizations of our experiment. However, this procedure is *ad hoc* and a better approach may exist.

**Benchmark Models for Comparison**

Supplementary Table 1 describes 5 variants of FEAT that we benchmarked in order to validate the algorithmic changes proposed above. We conducted this experiment to test the following hypotheses: 1) restricting FEAT to boolean operators would produce simpler models; 2) the post-run simplification operator would produce simpler models; 3) post-run simplification would produce models with derived features that were more orthogonal; 4) the multi-dimensional architecture FEAT uses would perform better than an even simpler "single model" approach frequently used in genetic programming.

In order to test these changes generally, we chose a set of 20 benchmark classification problems from the Penn ML Benchmark (PMLB).[22] These datasets are widely available, real-world and simulated problems. We chose 20 datasets whose shape (number of samples and features) was closest to that of the hypertension problems (Supplementary Table 2). For the PMLB comparisons, we ran 10 trials of shuffled 75/25 train/test splits.

#### Association between laboratory results and medications

To understand the maximum calcium feature that FEAT learned to classify apparent treatment-resistant hypertension, we performed multivariate logistic regression considering all anti-hypertensive medication features using backwards selection, optimizing for Bayesian Information Content.

### EXTENDED RESULTS

#### FEAT Method Benchmark

Supplementary Figure 1 displays performance comparisons of the FEAT variants (listed in Supplementary Table 1) on the 20 PMLB benchmark tasks. As shown in Supplementary Figure 1 (left), there were insignificant differences in AUPRC between methods except for Feat_1dim, which showed lower discrimination ($p$ <= 1e-3). However, the modifications to FEAT (simplification, Boolean operators and single dimensionality) all resulted in successively smaller model sizes (Supplementary Fig. 1, right). FEAT_boolean_simplify produced the smallest models across FEAT variants without clear drop in predictive performance. We considered restricting FEAT to produce models with only a single derived feature (Feat_1dim), but found that while it further decreased median model size by 71% ($p$=1.4x10$^{-18}$) it also decreased average precision by 4.1% ($p$=1.5x10$^{-4}$). Therefore Feat_boolean_simplify was used as the FEAT configuration for subsequent applications for computable phenotyping.

**Model Interpretability**

Supplementary Figure 3 shows the SHAP values generated for the random forest model for aTRH. Supplementary Figure 3A shows that expected risk factors for aTRH were important predictions in the RF model. For example, the most important feature, *low skewness* in the number of encounters per year while prescribed three or more hypertension medications ("# enc on 3 meds, skewness"), has a large positive impact on the model output. In other words, patients with a high number of such encounters in most years and a low number of such encounters in a minority of years (i.e. negatively skewed distribution) were more likely to be predicted as aTRH. A similar analysis can be extended to all 331 features incorporated in the model, although doing so is difficult given how many features conferred a non-negligible impact on the model.

The random forest model decision plot (Supplementary Fig. 3B) illustrates the impact of individual features on individual predictions; the top 20 most important features are shown. This plot depicts similar complexity to that of the LR L1 model (Figure 6), with a slow decay in importance across features. Thus, one cannot simply identify specific factors that explain classifications. For example, there are many features that appear to have had small, positive impacts resulting in misclassification of the single depicted false-positive subject (Supplementary Fig 3B, dot-dashed line with model output probability greater than 0.5). The mechanism by which each feature contributes to the misclassification cannot be deduced without fully considering the interactions between features in the ensemble. In contrast, since FEAT performs logistic regression on the transformed features (Figure 6), the derived predictors have linear and additive impacts on model output that can explain misclassifications.

For the regression models LR L1 and FEAT, two sets of SHAP values are estimated. The alternative approach of SHAP value estimation for aTRH models is provided in Supplementary Figure 4. In contrast to Figure 6, the SHAP values in Supplementary Figure 4 consider interactions amongst input data when estimating importance. In this case, SHAP values do not explicitly represent linear model coefficients. Instead, SHAP values are transformed by applying a linear projection to the input data and model coefficients. Put simply, whereas Figure 6 is faithful to the models and its coefficients, Supplementary Figure 4 shows feature importance estimates that are more faithful to the correlation structure of the input data. We note that considering such correlations gives a much more intuitive interpretation of the LR L1 model's important predictors, including small positive effects on aTRH predictions from encounter counts while prescribed multiple medications, systolic blood pressure summarizations, and counts of days on hypertension medications. It is worth noting that to correctly identify the relationships between such features and the LR L1 model predictions requires a close inspection of the data, and is not apparent from simple inspection of the model coefficients themselves (i.e. Figure 6). In contrast, in accounting for data collinearity in interpreting the FEAT model, while we do observe some smearing of the features' apparent impact the overall interpretability and interpretation of the model does not fundamentally change (Supplementary Fig. 4C).

### Clinical Chart Review

Patients were deemed to have hypertension if they had multiple documented elevated blood pressure measurements (SBP >= 140 mmHg or DBP >= 90), were being treated with an anti-hypertensive medication for blood pressure control, or had documented hypertension in diagnosis codes or notes. Elevated blood pressures were considered not indicative of hypertension if there was no clinical diagnosis and the elevation was potentially explained by clinical context, such as acute illness or pain, or interpreted as situational (e.g. white coat hypertension) and not treated as hypertension.

Patients were considered to have hypokalemia if there was documented evidence of an outpatient laboratory test result with low potassium or were prescribed outpatient oral potassium supplementation. Hypokalemia was considered explained if the measurements coincided with a dilutional explanation (e.g. saline infusion, chemo-infusion), acute illness potentially explaining (e.g. gastroenteritis with vomiting and diarrhea), dietary restriction, medication with known side effect (e.g. Bortezomib, amphotericin B), or hypomagnesemia.

Patients were considered to have apparent treatment-resistant hypertension (aTRH) if they were on anti-hypertension medications from 4 distinct classes for at least a month or from 3 distinct classes for over a month and had multiple elevated blood pressure measurements that did not appear to be explained by identifiable factors (e.g. medication adherence, insufficient dosing, acute illness). Patients with evidence of heart failure or chronic kidney disease prior to meeting aTRH criteria were considered negative.

16. La Cava W, Moore JH. Learning concise representations for regression by evolving networks of trees. In: *ICLR*. ICLR. ; 2019.

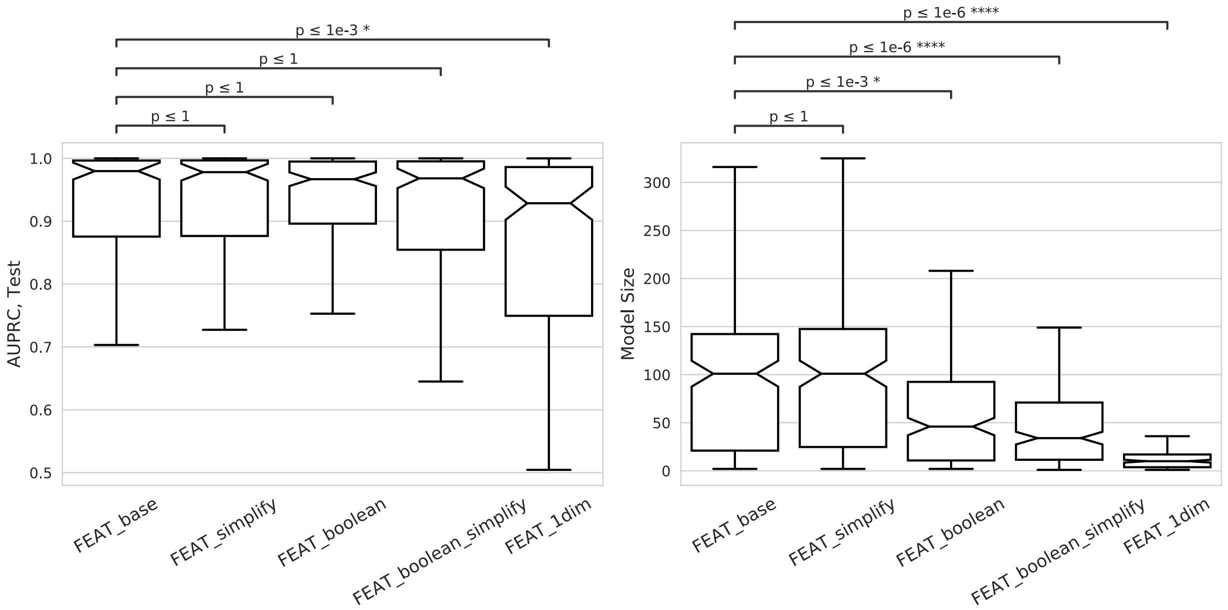

**Supplementary Figure 1: Evaluation of FEAT modifications.** *(Left)* Test AUPRC and *(Right)* model sizes of FEAT variants on 20 PMLB benchmark classification problems. Boxplots represent distribution of the mean 5-fold cross-validation test scores over 50 repeat realizations of the experiment. p values according to a Wilcoxon rank-sum test.

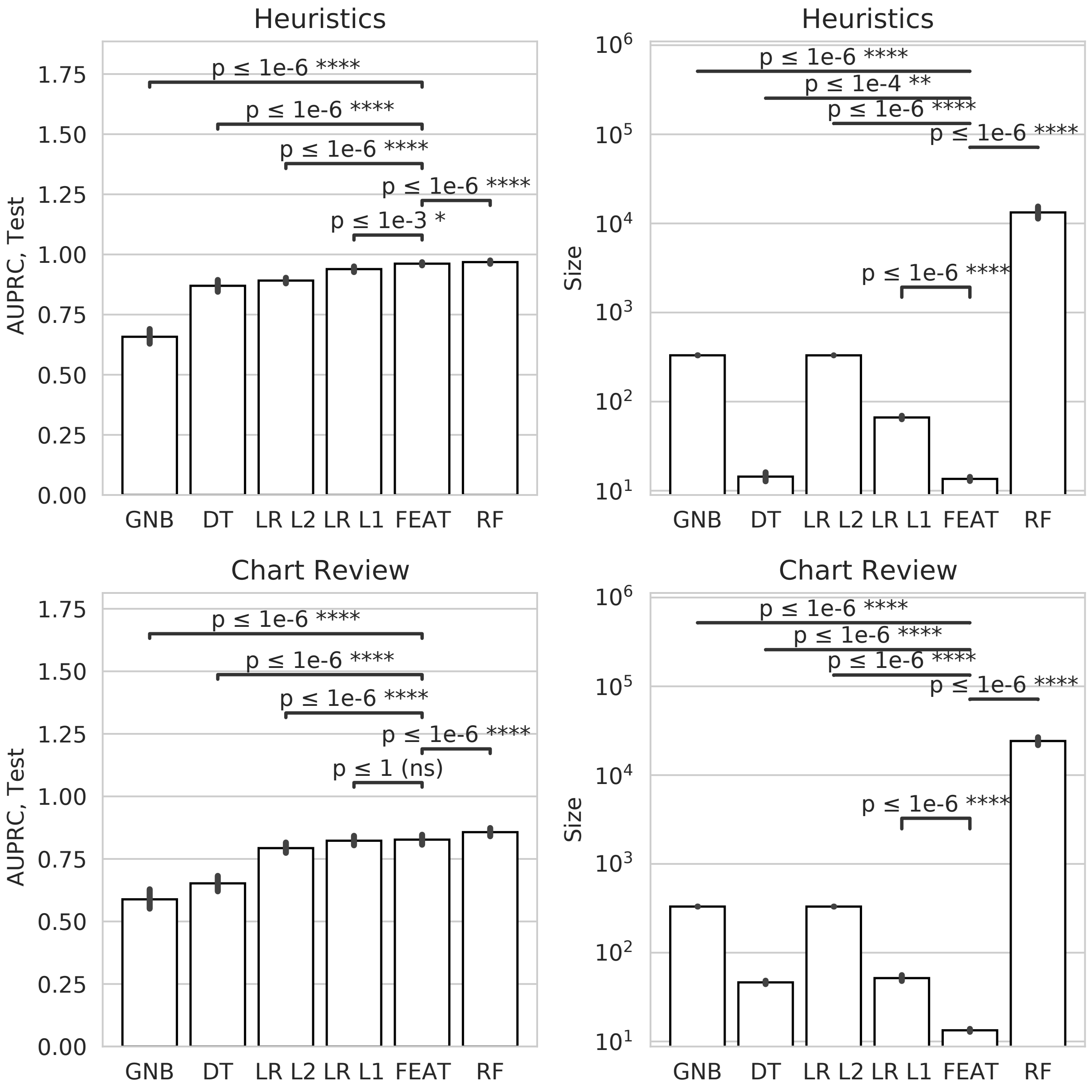

**Supplementary Figure 2: Comparison of discrimination and size of models trained to learn heuristics and chart-reviewed phenotypes**. Top plots indicate the rankings of methods according to AUPRC (left) and model size (right), when tasked with predicting the three expert heuristics. On the bottom, equivalent plots are shown for predicting the chart-reviewed phenotypes. Performance is ranked according to mean 5-fold CV performance and error bars indicate the standard error over 50 realizations of the experiment. $p$ values are calculated according to pairwise Wilcoxon rank-sum tests, with $\alpha = 0.001$.

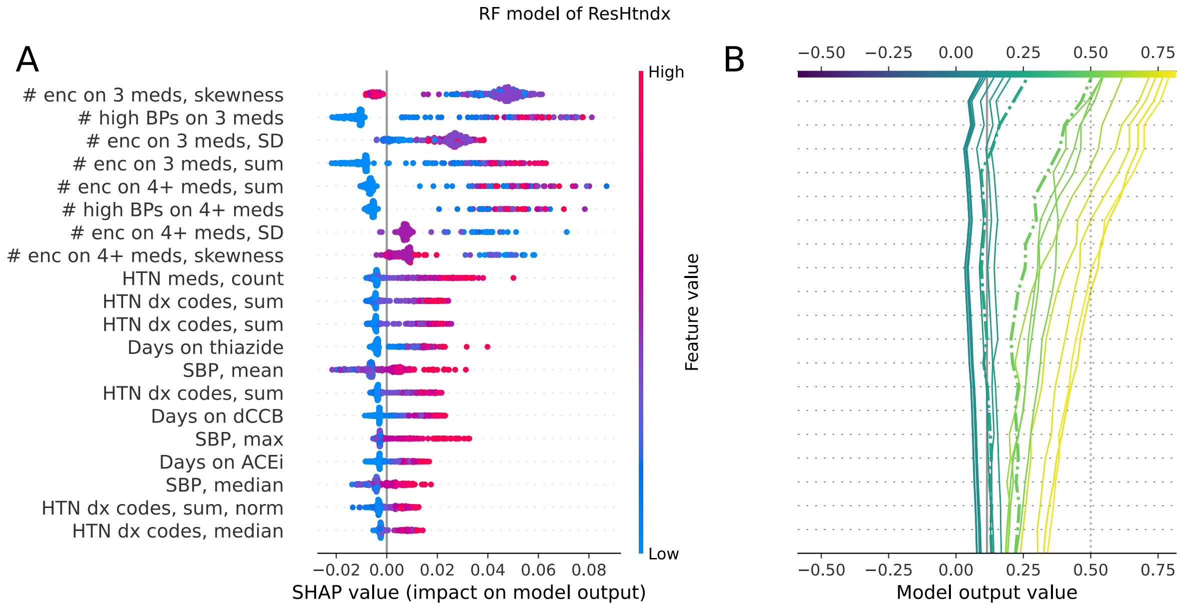

**Supplementary Figure 3**: **SHAP Plots for Random Forest Model Trained to Predict aTRH.** SHAP summary plots (left) and decision plots (right). The left plot indicates the most important features, ranked by the mean absolute SHAP value calculated on test data. The decision plot shows a sample of 10 positive and 10 negative point predictions by the models, with dotted lines indicating misclassifications.

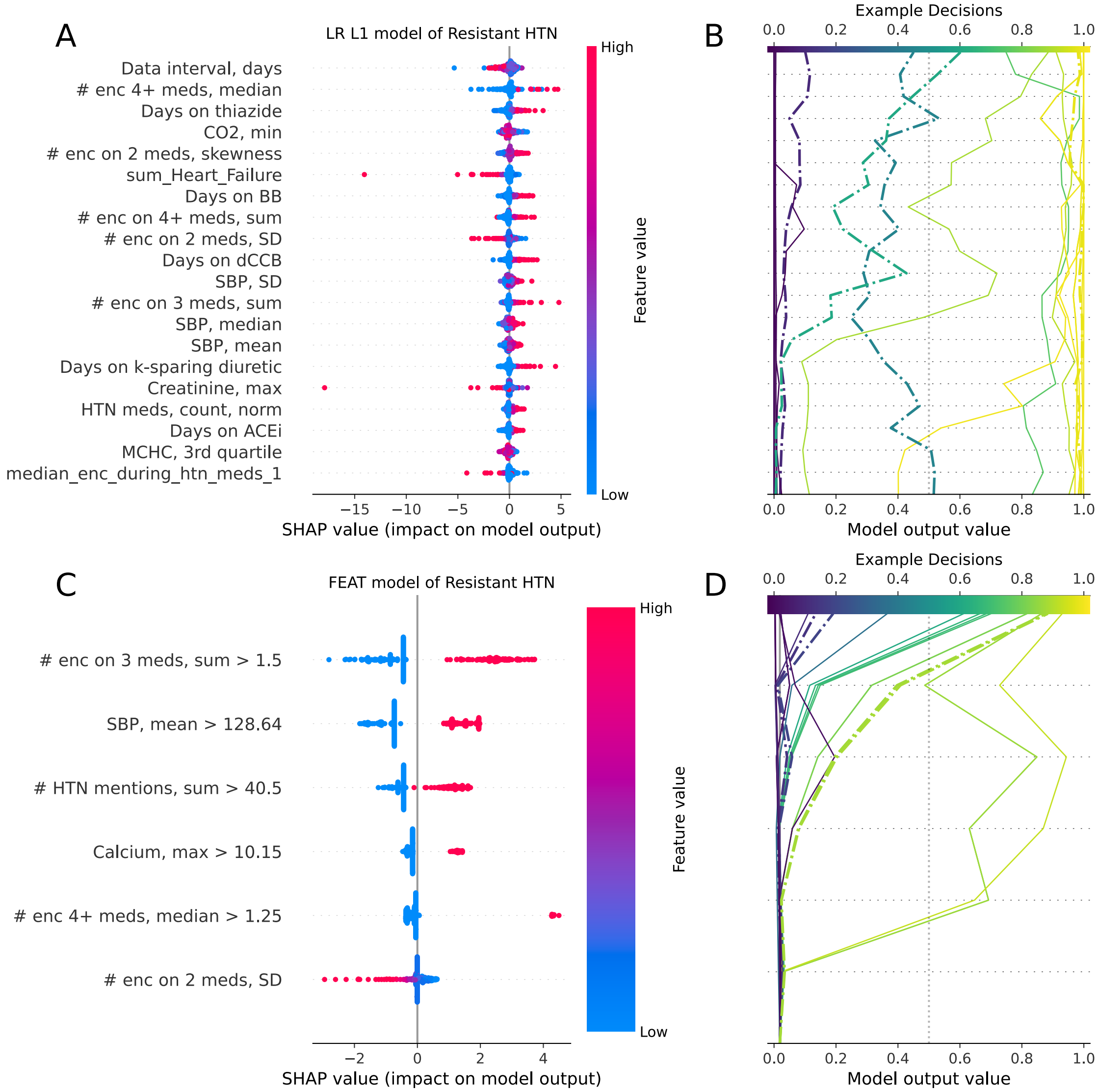

**Supplementary Figure 4**: **SHAP plots for LR L1 and FEAT Models Trained to Predict aTRH.** SHAP summary plots (A, C) and decision plots (B,D) according to the learned features of the LR L1 and FEAT models. In this case, SHAP values do not explicitly represent linear model coefficients. Instead, SHAP values are transformed by applying a linear projection to the input data and model coefficients, leading to feature importance estimates that are more faithful to the data than the model. The left plot indicates the most important features, ranked by the mean absolute SHAP value calculated on test data. The decision plot shows a sample of 10 positive and 10 negative point predictions by the models, with dotted lines indicating misclassifications.

**Supplementary Table 1:** FEAT method variants tested in benchmark experiment.

| Common settings | population size = 500, generations = 200, max_time = 1 hour |
|---|---|
| Feat_base | FEAT with default arguments |
| Feat_simplify* | FEAT with post-run simplification |
| Feat_boolean | FEAT restricted to Boolean operators |
| Feat_boolean_simplify | FEAT restricted to Boolean operators with post-run simplification |
| Feat_1dim | FEAT restricted to producing a single feature (one dimensional) |

**Supplementary Table 2:** Datasets from PMLB[22] used for benchmark comparisons.

| **Dataset Name** | **Number of Features** | **Number of Instances** |
|---|---|---|
| Hill_Valley_with_noise | 100 | 1212 |
| Hill_Valley_without_noise | 100 | 1212 |
| backache | 32 | 180 |
| breast-cancer-wisconsin | 30 | 569 |
| chess | 36 | 3196 |
| clean1 | 168 | 476 |
| clean2 | 168 | 6598 |
| coil2000 | 85 | 9822 |
| colic | 22 | 368 |
| dis | 29 | 3772 |
| horse-colic | 22 | 368 |
| hypothyroid | 25 | 3163 |
| ionosphere | 34 | 351 |
| kr-vs-kp | 36 | 3196 |
| molecular-biology_promoters | 58 | 106 |
| sonar | 60 | 208 |
| spambase | 57 | 4601 |
| spectf | 44 | 349 |
| tokyo1 | 44 | 959 |
| wdbc | 30 | 569 |

**Supplementary Table 3:** EHR features considered as potential predictors

| Group | VARIABLE | VARIABLE DESCRIPTION |
|---|---|---|
| Identifiers | UNI_ID | the unique study-generated identifier for Patient |
| Demo | age | patient's age at right-censoring date |
| | Male | patient's indicated Sex (1 = male, 0 = female) |
| | BLACK | 1 = black, 0 = non-black |
| | OTHER | 1 = asian, other, mixed,native american,pacific islander, 0 = black or white |
| | WHITE | 1 = white, 0 = non-white |
| | ZIP_CAT | distance from patient's home to 19104, in category |
| Encounter | MASTER_LOCATION_CODE | code for healthcare site (not one hot encoded), common service for all source systems. This is used to map UPHS's various versions of the same or similar codes into a matched list of services. This data will persist to the MDM level. |
| | GENERAL_INTERNAL_MEDICINE | 1 = IM practice, 0 = FM practice |
| BMI/Weight | weight_min/max/median/sd/skewness | min/max/median/sd/skewness of weights |
| | bmi_min/max/sd/skewness | min/max/sd/skewness of BMI |
| BP | bp_n | total number of bp measurements |
| | min_systolic | minimum of systolic blood pressure measured |
| | min_diastolic | minimum of diastolic blood pressure measured |
| | max_systolic | maximum of systolic blood pressure measured |
| | max_systolic | maximum of diastolic blood pressure measured |
| | mean_systolic | mean of systolic blood pressure measured |
| | mean_diastolic | mean of diastolic blood pressure measured |
| | median_systolic | median of systolic blood pressure measured |
| | median_diastolic | median of diastolic blood pressure measured |
| | sd_systolic | standard deviation of systolic blood pressure measured |
| | sd_diastolic | standard deviation of diastolic blood pressure measured |
| | skew_systolic | skewness of systolic blood pressure measured |
| | skew_diastolic | skewness of diastolic blood pressure measured |
| | high_bp_n | number of high blood pressure, SBP >= 140 or DBP >= 90 |
| | mean_high_bp_systolic | mean systolic bp of all high blood pressure measurements (SBP >=140 or DBP >=90) |
| | mean_high_bp_diastolic | mean diastolic bp of all high blood pressure measurements (SBP >=140 or DBP >=90) |
| | median_high_bp_systolic | median systolic bp of all high blood pressure measurements (SBP >=140 or DBP >=90) |
| | median_high_bp_diastolic | median diastolic bp of all high blood pressure measurements (SBP >=140 or DBP >=90) |
| | sd_high_bp_systolic | standard deviation of systolic bp of all high blood pressure measurements (SBP >=140 or DBP >=90) |
| | sd_high_bp_diastolic | standard deviation of diastolic bp of all high blood pressure measurements (SBP >=140 or DBP >=90) |
| | skew_high_bp_systolic | skewness of systolic bp of all high blood pressure measurements (SBP >=140 or DBP >=90) |
| | skew_high_bp_diastolic | skewness of diastolic bp of all high blood pressure measurements (SBP >=140 or DBP >=90) |
| | median/sd/skew_high_bp_n_yr | median/sd/skewness of high blood pressure measurements (SBP >=140 or DBP >=90) per year |
| Labs | max.lab_XXX | maximum of XXX lab test |
| | min.lab_XXX | minimum of XXX lab test |

| | | |
|---|---|---|
| | median.lab_XXX | median of XXX lab test |
| | q1.lab_XXX | 1st quantile of XXX lab test |
| | q3.lab_XXX | 3rd quantile of XXX lab test |
| **Dx** | median_ICD_XXX (Dx) | median XXX ICD-9 and ICD-10 codes, by year |
| | sum_ICD_XXX (Dx) | sum XXX ICD-9 and ICD-10 codes, by year |
| | median_XXX (disease name) | median XXX disease name, by year |
| | sum_XXX (disease name) | sum XXX disease name, year |
| | Dx_N | number of total ICD-9 and ICD-10 codes (PK_DX_ID) |
| | enc_N | number of OUTPATIENT (including INFUSION VISIT) encounters |
| | dx_days_x | days from 1st Dx to last Dx in system |
| **Medication** | HTN_MED_days_XXX | days on med XXX (including anti-HTN and Potassium Supplement) |
| | MED_N | number of medication prescriptions total |
| | high_BP_during_htn_meds_1/2/3/4_plus | number of high BP measurements during 1/2/3/4+ anti-HTN meds |
| | sum_enc_during_htn_meds_1/2/3/4_plus | number of OUTPATIENT encounters during 1/2/3/4+ meds |
| | median_enc_during_htn_meds_1/2/3/4_plus | median number (by year) of OUTPATIENT encounters during 1/2/3/4+ meds |
| | sd_enc_during_htn_meds_1/2/3/4_plus | sd of number (by year) of OUTPATIENT encounters during 1/2/3/4+ meds |
| | skewness_enc_during_htn_meds_1/2/3/4_plus | skewness of number (by year) of OUTPATIENT encounters during 1/2/3/4+ meds |
| | N_med_K_chlo_enc | number of encounters on POTASSIUM_CHLORIDE/POTASSIUM_GLUCONATE |
| | sd_med_K_chlo_enc | sd of number (by year) of encounters on POTASSIUM_CHLORIDE/POTASSIUM_GLUCONATE |
| | skewness_med_K_chlo_enc | skewness of number (by year) of encounters on POTASSIUM_CHLORIDE/POTASSIUM_GLUCONATE |
| **Heuristic Features** | low_K_N | # of low potassium test results |
| | test_K_N | # of potassium test results |
| | Med_Potassium_N | # of potassium supplement medication subscriptions |
| | Dx_HypoK_N | # of Hypokalemia Dx |
| **HTN Score Features** | ICD_hyp_sum | HTN ICD codes |
| | MED_HTN_N | anti-HTN med prescriptions |
| | bp_hyp_norm | high_bp_n/bp_n |
| | ICD_hyp_sum_norm | ICD_hyp_sum/Dx_N |
| | MED_HTN_N_norm | MED_HTN_N/MED_N |
| | re_hyp_spe_norm | re_htn_spec/words_n |
| **Regex** | re_htn_sum | sum of regex counts in clinical notes for hypertension |
| | re_htn_spec_sum | sum of regex counts in clinical notes for hypertension (specific, excluding preliminary negations) |
| | re_htn_teixera_sum | sum of regex counts in clinical notes for hypertension (regex used in Teixeira paper) |
| | re_word_count_sum | sum word counts in clinical notes |
| | re_htn_max | maximum of regex counts in clinical notes for hypertension |
| | re_htn_spec_max | maximum of regex counts in clinical notes for hypertension (specific, excluding preliminary negations) |
| | re_htn_teixera_max | maximum of regex counts in clinical notes for hypertension (regex used in Teixeira paper) |
| | re_word_count_max | maximum word counts in clinical notes |
| | re_htn_mean | mean of regex counts in clinical notes for hypertension |

| | | |
|---|---|---|
| | re_htn_spec_mean | mean of regex counts in clinical notes for hypertension (specific, excluding preliminary negations) |
| | re_htn_teixera_mean | mean of regex counts in clinical notes for hypertension (regex used in Teixeira paper) |
| | re_word_count_mean | mean word counts in clinical notes |
| | re_htn_median | median of regex counts in clinical notes for hypertension |
| | re_htn_spec_median | median of regex counts in clinical notes for hypertension (specific, excluding preliminary negations) |
| | re_htn_teixera_median | median of regex counts in clinical notes for hypertension (regex used in Teixeira paper) |
| | re_word_count_median | median word counts in clinical notes |
| | re_htn_sd | standard deviation of regex counts in clinical notes for hypertension |
| | re_htn_spec_sd | standard deviation of regex counts in clinical notes for hypertension (specific, excluding preliminary negations) |
| | re_htn_teixera_sd | standard deviation of regex counts in clinical notes for hypertension (regex used in Teixeira paper) |
| | re_word_count_sd | standard deviation of word counts in clinical notes |
| | re_htn_skewness | skewness of regex counts in clinical notes for hypertension |
| | re_htn_spec_skewness | skewness of regex counts in clinical notes for hypertension (specific, excluding preliminary negations) |
| | re_htn_teixera_skewness | skewness of regex counts in clinical notes for hypertension (regex used in Teixeira paper) |
| | re_word_count_skewness | skewness of word counts in clinical notes |

**Supplementary Table 4:** EHR laboratory results considered as predictors

| Labs |
|---|
| Pct.BASOPHILS |
| Pct.EOSINOPHILS |
| Pct.LYMPHOCYTES |
| Pct.MONOCYTES |
| Pct.NEUTROPHILS |
| ALBUMIN |
| ALKALINE.PHOSPHATASE |
| ALT |
| AST |
| BILIRUBIN.TOTAL |
| CALCIUM |
| CARBON.DIOXIDE |
| CHLORIDE |
| CHOLESTEROL |
| CHOLESTEROL.CALCULATED.LOW.DENSITY.LIPOPROTEIN |
| CHOLESTEROL.CALCULATED.HIGH.DENSITY.LIPOPROTEIN |
| CREATININE |
| HEMATOCRIT |
| HEMOGLOBIN |
| MEAN.CELLULAR.HEMOGLOBIN |
| MEAN.CELLULAR.HEMOGLOBIN.CONCENTRATION |
| MEAN.CELLULAR.VOLUME |
| PLATELETS |
| POTASSIUM |
| PROTEIN.TOTAL |
| RDW |
| RED.BLOOD.CELLS |
| SODIUM |
| THYROID.STIMULATING.HORMONE |
| TRIGLYCERIDES |
| UREA.NITROGEN |
| WBC |

**Supplementary Table 5:** EHR diagnosis codes considered as predictors, encoded as median count per year

| **median_ICD_XXX (Dx)** |
|---|
| median_E03_9 |
| median_E11_9 |
| median_E78_00 |
| median_E78_01 |
| median_E78_2 |
| median_E78_5 |
| median_I10 |
| median_I16_0 |
| median_I16_1 |
| median_I16_9 |
| **median_XXX (disease name)** |
| median_Diabetes_type_1 |
| median_Dyslipidemias |
| median_Essential_HTN |
| median_HTN_Emergency |
| median_Hypothyroidism |

**Supplementary Table 6:** EHR diagnosis codes considered as predictors, encoded as total count

| **sum_ICD_XXX (Dx)** |
|---|
| sum_E03_8 |
| sum_E03_9 |
| sum_E11_65 |
| sum_E11_9 |
| sum_E66_01 |
| sum_E66_09 |
| sum_E66_1 |
| sum_E66_8 |
| sum_E66_9 |
| sum_E78_00 |
| sum_E78_01 |
| sum_E78_2 |
| sum_E78_5 |
| sum_E87_6 |
| sum_G47_30 |
| sum_G47_33 |
| sum_I10 |
| sum_I16_0 |
| sum_I16_1 |
| sum_I16_9 |
| sum_I25_10 |
| sum_I48_0 |
| sum_I48_1 |
| sum_I48_2 |
| sum_I48_91 |
| sum_L70_8 |
| sum_N18_3 |
| **sum_XXX (disease name)** |
| sum_ACNE |
| sum_Arrythmias |
| sum_Atrial_fibrillation |
| sum_CAD_native |
| sum_CKD |
| sum_Diabetes_type_2 |
| sum_Dyslipidemias |

| |
|---|
| sum_Essential_HTN |
| sum_Heart_Failure |
| sum_HTN_Emergency |
| sum_Hypokalemia |
| sum_Hypothyroidism |
| sum_Obesity |
| sum_Obstructive_Sleep_Apnea |

**Supplementary Table 7:** Anti-hypertensive medication features considered, encoded as number of days prescribed

| **HTN_MED_days_XXX** |
|---|
| HTN_MED_days_ACEI_ARB |
| HTN_MED_days_ALDOSTERONE_ANTAGONIST |
| HTN_MED_days_ALDOSTERONE_ANTAGONISTS |
| HTN_MED_days_ALPHA_ANTAGONISTS |
| HTN_MED_days_BETA_BLOCKERS |
| HTN_MED_days_CENTRAL_ALPHA_AGONISTS |
| HTN_MED_days_DIHYDRO_CCBS |
| HTN_MED_days_HYDRALAZINE |
| HTN_MED_days_K_SPARING_DIURETICS |
| HTN_MED_days_LOOP_DIURETICS |
| HTN_MED_days_MINOXIDIL |
| HTN_MED_days_NON_DIHYDRO_CCBS |
| HTN_MED_days_RENIN_ANTAGONIST |
| HTN_MED_days_THIAZIDE |
| HTN_MED_days_POTASSIUM_CHLORIDE |

**Supplementary Table 8:** EHR features included in trained computable phenotypes

| Full Name | Short Name |
|---|---|
| **Days between 1st dx to last dx code** | Data interval, days |
| **Number of high BP measurements while on 3 anti-HTN meds** | # high BPs on 3 meds |
| **Number of high BP measurements while on 4+ anti-HTN meds** | # high BPs on 4+ meds |
| **Days prescribed ACE inhibitors** | Days on ACEi |
| **Days prescribed beta blockers** | Days on BB |
| **Days prescribed dihydropyridine calcium channel blockers** | Days on dCCB |
| **Days prescribed potassium sparing diuretics** | Days on k-sparing diuretic |
| **Days prescribed thiazides** | Days on thiazide |
| **Sum of HTN ICD codes** | HTN dx codes, sum |
| **Sum of HTN ICD codes divided by the total number of ICD codes** | HTN dx codes, sum, norm |
| **Maximum of systolic blood pressure measured** | SBP, max |
| **Maximum of calcium measured** | Calcium, max |
| **Maximum of creatinine measured** | Creatinine, max |
| **Mean of systolic blood pressure measured** | SBP, mean |
| **Number of anti-hypertension medication prescriptions** | HTN meds, count |
| **Number of anti-hypertension medication prescriptions divided by the total number of prescribed medications** | HTN meds, count, norm |
| **Median number (by year) of OUTPATIENT encounters during 4+ anti-hypertension medications** | # enc 4+ meds, median |
| **Count of I10 (hypertension) ICD codes, median per year** | HTN dx codes, median |
| **Median of systolic blood pressure measured** | SBP, median |
| **Median of potassium measured** | K, median |
| **Minimum of potassium measured** | K, min |
| **Sum of regex counts in clinical notes for hypertension** | # HTN mentions, sum |
| **Standard deviation of number (by year) of OUTPATIENT encounters during 2 anti-hypertension medications** | # enc on 2 meds, SD |
| **Standard deviation of number (by year) of OUTPATIENT encounters during 3 anti-hypertension medications** | # enc on 3 meds, SD |
| **Standard deviation of number (by year) of OUTPATIENT encounters during 4+ anti-hypertension medications** | # enc on 4+ meds, SD |
| **Standard deviation of systolic blood pressure measured** | SBP, SD |
| **Skewness of number (by year) of OUTPATIENT encounters during 2 anti-hypertension medications** | # enc on 2 meds, skewness |

| | |
|---|---|
| **Skewness of number (by year) of OUTPATIENT encounters during 3 anti-hypertension medications** | # enc on 3 meds, skewness |
| **Skewness of number (by year) of OUTPATIENT encounters during 4+ anti-hypertension medications** | # enc on 4+ meds, skewness |
| **Sum of number (by year) of OUTPATIENT encounters during 3 anti-hypertension medications** | # enc on 3 meds, sum |
| **Sum of number (by year) of OUTPATIENT encounters during 4+ anti-hypertension medications** | # enc on 4+ meds, sum |
| **Sum of I10 (hypertension) ICD codes** | HTN dx codes, sum |
| **Sum of I10 (hypertension) ICD codes** | HTN dx codes, sum |
| **High Density Lipoprotein (HDL) cholesterol, min** | HDLc, min |
| **High Density Lipoprotein (HDL) cholesterol, max** | HDLc, max |
| **Chloride, 1st quartile** | Chloride, min |
| **Carbon dioxide, min** | CO2, min |
| **Mean cellular hemoglobin concentration (MCHC), 3rd quartile** | MCHC, 3rd quartile |
| **Platelets, max** | Platelets, max |
| **Mean cellular hemoglobin concentration (MCHC), 1st quartile** | MCHC, 1st quartile |
| **Low Density Lipoprotein (LDL) cholesterol, calculated, median** | LDLc, median |
| **Regex counts in clinical notes for hypertension, median per year** | # of HTN mentions, median |
| **Albumin, min** | Albumin, min |
| **Calcium, 1st quartile** | Calcium, 1st quartile |
| **Chloride, median** | Chloride, median |
| **Thyroid stimulating hormone (TSH), 1st quartile** | TSH, 1st quartile |
